# Diffusion Deep Learning for Brain Age Prediction and Longitudinal Tracking in Children Through Adulthood

**DOI:** 10.1101/2023.10.17.23297166

**Authors:** Anna Zapaishchykova, Divyanshu Tak, Zezhong Ye, Kevin X. Liu, Jirapat Likitlersuang, Sridhar Vajapeyam, Rishi B. Chopra, Jakob Seidlitz, Richard AI Bethlehem, Lifespan Brain Chart Consortium, Raymond H. Mak, Sabine Mueller, Daphne A. Haas-Kogan, Tina Y. Poussaint, Hugo J.W.L. Aerts, Benjamin H. Kann

## Abstract

Deep learning (DL)-based prediction of biological age in the developing human from a brain magnetic resonance image (MRI) (“*brain age*”) may have important diagnostic and therapeutic applications as a non-invasive biomarker of brain health, aging, and neurocognition. While previous deep learning tools for predicting brain age have shown promising capabilities using single-institution, cross-sectional datasets, our work aims to advance the field by leveraging multi-site, longitudinal data with externally validated and independently implementable code to facilitate clinical translation and utility. This builds on prior foundational efforts in brain age modeling to enable broader generalization and individual’s longitudinal brain development. Here, we leveraged 32,851 T1-weighted MRI scans from healthy children and adolescents aged 3 to 30 from 16 multisite datasets to develop and evaluate several DL brain age frameworks, including a novel regression diffusion DL network (AgeDiffuse). In a multisite external validation (5 datasets), we found that AgeDiffuse outperformed conventional DL frameworks, with a mean absolute error (MAE) of 2.78 years (IQR:[1.2-3.9]). In a second, separate external validation (3 datasets), AgeDiffuse yielded an MAE of 1.97 years (IQR: [0.8-2.8]). We found that AgeDiffuse brain age predictions reflected age-related brain structure volume changes better than biological age (R2=0.48 vs R2=0.37). Finally, we found that longitudinal predicted brain age tracked closely with chronological age at the individual level. To enable independent validation and application, we made AgeDiffuse publicly available and usable for the research community.

**Highlights:**

- Diffusion regression models trained with a large dataset (AgeDiffuse) enable accurate pediatric brain age prediction.
- AgeDiffuse demonstrates relatively stable performance on multiple external validation sets across people aged 3 – 30.
- Our pipeline is made publicly accessible, encouraging collaboration and progress in pediatric brain research.

## 1. Introduction

The prediction of biological age from healthy brain magnetic resonance imaging (MRI) scans (i.e. “*brain age*”) has the potential for wide-ranging medical and scientific applications ^1,2^. Establishing reliable brain age prediction in large healthy-control populations would enable studying how various diseases, interventions, and socioeconomic factors influence brain development. When examined within cohorts affected by particular risk factors, the difference between predicted brain age and actual chronological age (i.e. “*brain age gap*”) may yield insights into how various external and internal factors affect brain development ^3,4^. Increased brain age gap has been associated with several brain disorders, such as schizophrenia, multiple sclerosis, mild cognitive impairment, and dementia ^5^. Furthermore, accurately tracking the brain age gap may be useful in evaluating therapies designed to prevent neurocognitive disorder. Most research to this point has centered on adult and elderly conditions, where accelerated brain aging is inherently seen as a negative factor ^6^. The implications of the brain age gap in developing children and young adults remain unclear, mainly owing to a lack of robust models that can accurately predict brain age out-of-sample ^7^. The existing brain age prediction models have limited generalizability because they fail to make accurate predictions on new datasets that differ from the data used for model training ^8^.

Researchers have explored multiple approaches to brain age prediction, leading to a diverse set of methods with varying results ^9^. Direct comparison of these methods is challenging due to cross-study population differences, various imaging preprocessing techniques, and different evaluation strategies. Deep learning (DL) has emerged as a popular strategy for brain age prediction, given its remarkable success in trans-domain image analysis problems and its avoidance of time-consuming traditional feature extraction and preprocessing steps ^9^. Within pediatric or developing brain age prediction, there have been relatively few investigations ^10–13^, likely due to limited data availability in this age range. Most existing studies demonstrate their models on single-institution datasets and have lacked multi-institutional external validations ^10–13^, which is crucial for assessing true model generalization across diverse real-world settings and clinical utility. Factors including differences in scanners and protocols across sites, patient demographics, and other manifestations of dataset shift and drift are known to impact performance significantly ^14,15^. Furthermore, reviewing the pre-existing literature, we found no pediatric brain age models with implementable codes ^10–13^, which is critical to moving the field forward and investigating these models’ clinical utility ^16^. Finally, brain age models developed from cross-sectional data may not be suitable for individual brain age tracking, and further study is needed to determine how brain age models perform across longitudinal time points, and their relationship to structural brain changes ^17,18^.

In this study, we aim to address these gaps and develop a usable open-source model for reliable brain age prediction for childhood through young adulthood. Given the recent rise of generative DL methods and their promising results within the medical imaging domain ^19^, we developed a diffusion dual-guidance probabilistic regression model for pediatric brain age prediction (AgeDiffuse). We compared it to the state-of-the-art convolutional neural network (CNN) approaches, making this the first work, to our knowledge, to adapt diffusion models for image-based regression tasks. We demonstrate that diffusion-based models generalize well across two tiers of external validation, encompassing multi-institutional datasets from diverse geographic regions. We also investigate structural brain changes and their correlations with longitudinal brain age changes to yield interpretable insights into the model’s inner workings. Altogether, we present a robust model rigorously validated and made publicly available to the community, enabling the investigation of pediatric brain age in various clinical scenarios.

## 2. Results

### 2.1. Diffusion Regression for Brain Age

We aggregated a dataset with 32,851 MRI T1-weighted (T1w) scans (Train Set N=4,549, Test Set 1 N=583, Test Set 2 N=27,719) from subjects aged 3-30 years from 16 publicly available, multisite datasets of healthy, developing children through adulthood (Fig. 1A; Methods “Dataset” section). We then developed an MRI preprocessing and registration pipeline (Fig. 1B, see Methods “Image Preprocessing and **Registration**” section). We evaluated the performance of two state-of-the-art DL approaches for medical imaging: 1) a medical-domain, pretrained 2D convolutional neural network (RagImageNet^20^) and 2) a self-supervised, pretrained 3D UNet (ModelGenesis^21^) (see Supplementary Material A1. Model hyperparameter tuning). We then developed a 2D diffusion-based regression model, called AgeDiffuse model, that uses dual-granularity guidance and condition-specific maximum mean discrepancy (MMD) regularization. AgeDiffuse was adapted from a dual-guidance diffusion model for medical image classification ^19^ (see Methods “Regression Dual-Guidance Diffusion Model”). Dual-guidance models use use both global and local priors for conditional guidance at each step, and have the advantage of modeling representations with both holistic and fine-grained understanding of medical images.

**Figure 1.**
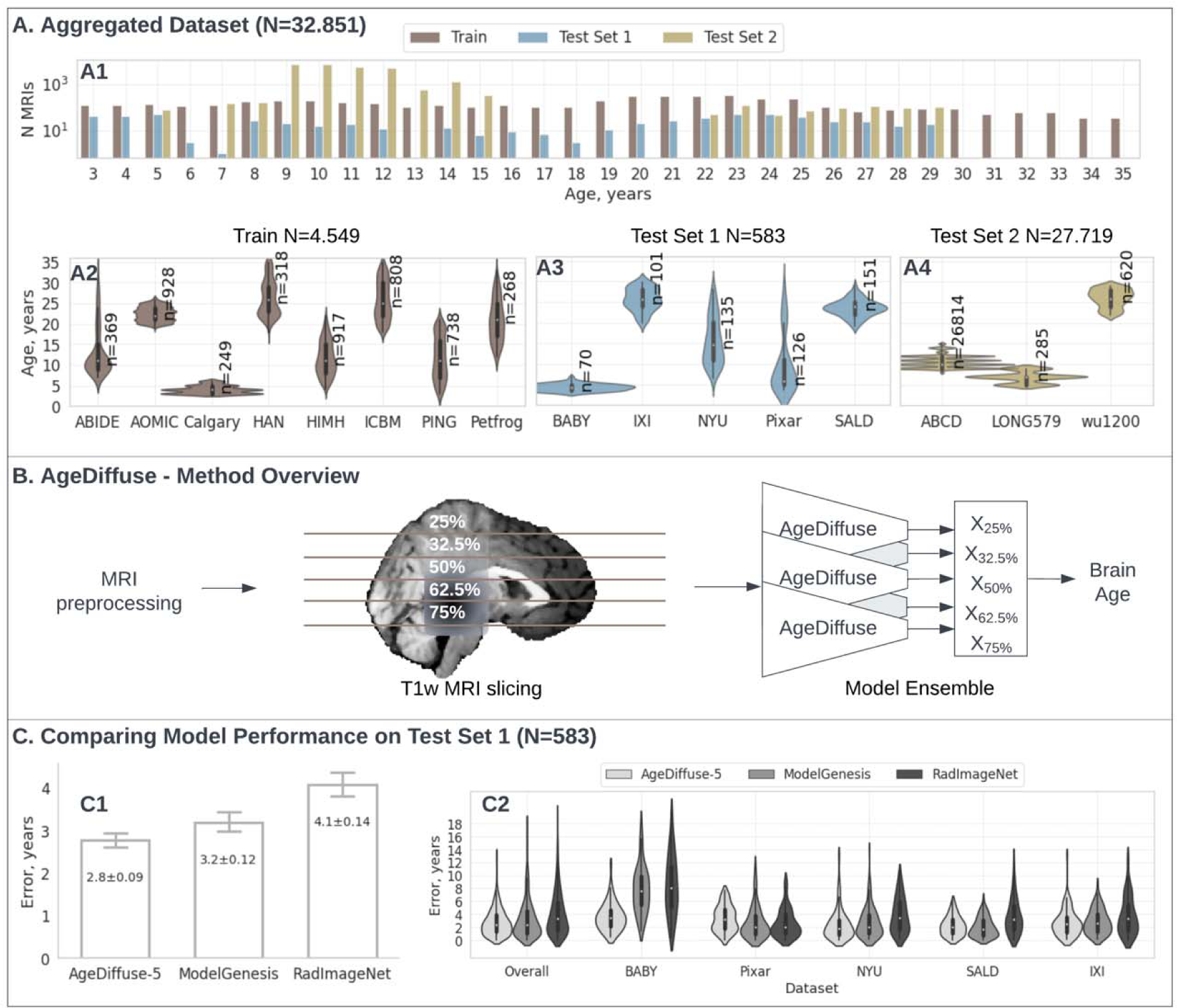
**A.** Aggregated dataset overview (total N=32,851). **A1:** Bar plot with number of MRI T1w per age group in Train (N=4,549)/Test Set 1(N=583)/Test Set 2(N=27,719); the y-axis is log scaled. **A2- A4**: Violin plots for dataset age distributions in Train(A2)/Test Set 1(A3)/Test Set 2(A4). The violins represent kernel density estimates of the age distribution in each dataset. Wider sections of the violins indicate a higher probability density at that error level . **B.** AgeDiffuse method overview: MRI preprocessing, 2D slice selection, AgeDiffuse model prediction, and model ensembling . **C.** Model performance comparisons on Test Set 1 (N=583; 5 datasets). **C1:** Bar plot for model-wise mean comparison in Test Set 1, with 95% confidence intervals overlay. The diffusion 5-slice ensemble (AgeDiffuse-5) performed with the highest accuracy among all models with mean error 2.8 years[IQR=1.3-3.9] compared to ModelGenesis mean error 3.2 years [IQR=1.0-4.5] and RadImageNet mean error 4.1 years [IQR=1.5-5.8]. **C2:** Violin plots for model-wise error distribution comparison in Test Set 1. MRI = Magnetic resonance imaging, AgeDiffuse = Novel regression dual- guidance diffusion model for brain age prediction.

On initial multi-institutional external validation (Test Set 1, N=583, 5 datasets), the diffusion network using the median axial slice as input (AgeDiffuse-1) achieved the highest accuracy compared to other methods for predicting chronological age (Table 1, MAE = 3.15 years, IQR=[1.27-4.41]Table *1*). To investigate if sampling from multiple axial slices would improve model performance, we trained 2D diffusion models on axial MRIs sampled from the 25, 27.5, 50 (median), 62.5, and 75 percentile slices in the craniocaudal distribution and then tested model ensembling across slices (see Methods “Model Ensembling”). The 5-slice diffusion network ensemble (AgeDiffuse-5) achieved the highest accuracy with MAE=2.78 years (IQR=[1.24-3.92]) outperforming 3D approach ModelGenesis MAE=3.19 years (IQR=[1.0-4.5]) and 2D RadImageNet MAE=4.07 years ([IQR=1.5-5.8]). To further test the model generalizability, we conducted a blinded secondary validation on three external datasets (Test Set 2; N=27,719). We compared simple model averaging with varying sizes and outlier exclusion to evaluate different ensembling techniques and found that the five-slice AgeDiffuse-5 model yielded the best brain age prediction with MAE=1.97 years (IQR=[0.76-2.75]) (Figure 2). For all models, accuracy decreased for later ages, particularly over 25 years old, though AgeDiffuse had less performance degradation than other models (See Supplementary Figure S6 and Supplementary A4. Outlier Analysis).

**Table 1.** Comparison of mean absolute error (MAE) between different models on Test Set 1. 2D equidistant quantile slices ensembling (AgeDiffuse-5) provides a robust prediction while being less susceptible to noise and outperforms other methods. IQR=interquartile range.

| Method | 2d/3d | MAE, years [IQR] |
| --- | --- | --- |
| RadImageNet | 2D – median slice | 4.07 [1.5-5.8] |
| ModelGenesis | 3D | 3.19 [1.0-4.5] |
| AgeDiffuse-1 | 2D – median slice | 3.15 [1.27-4.41] |
| AgeDiffuse-5 | 2D Model ensemble: 25th, 37.5th, median, 62.5th, 75th slices | 2.78 [1.24-3.92] |

**Figure 2.**
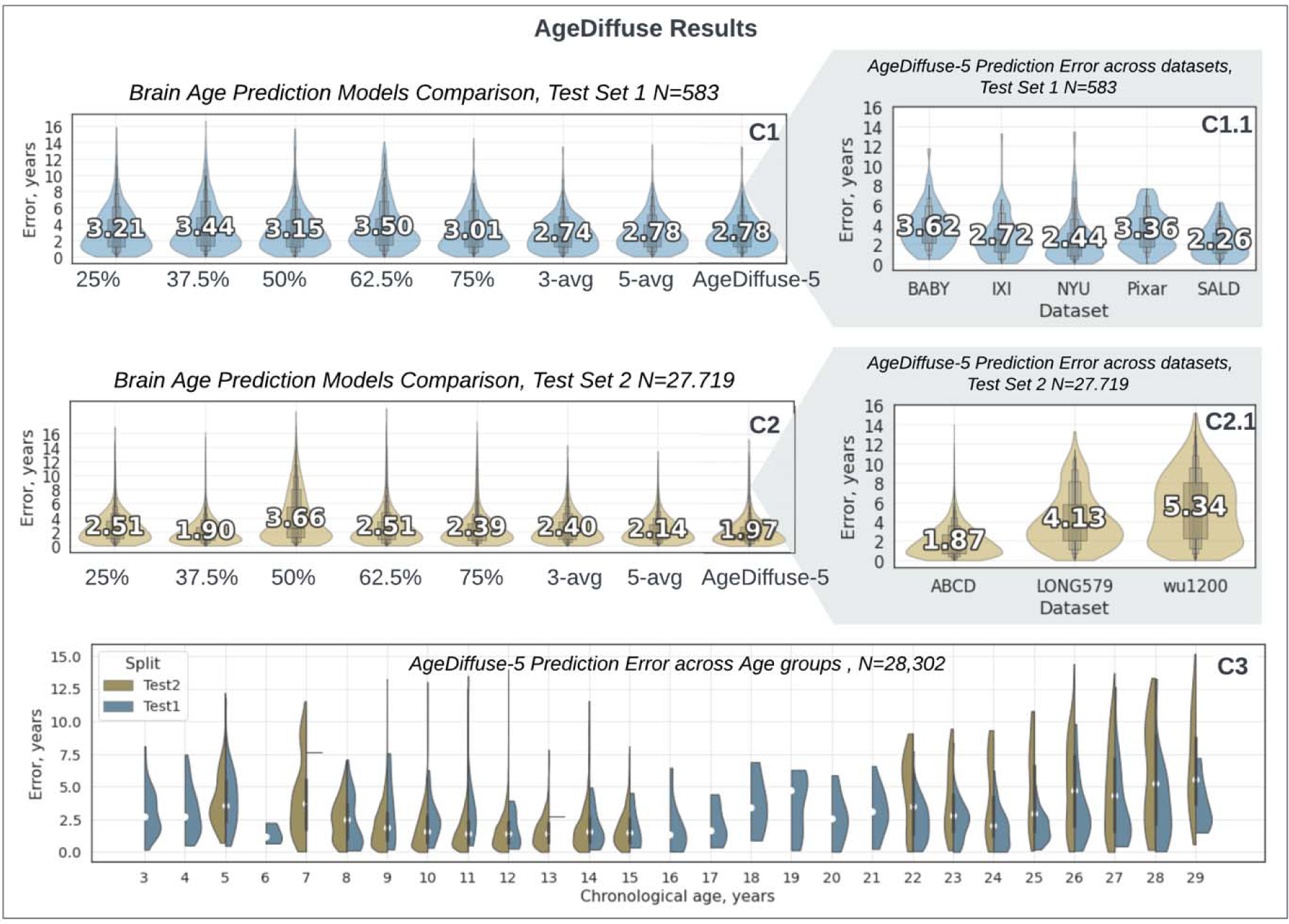
Violin plots for AgeDiffuse brain age prediction in developing children: dual-tiered external validation with text median overlays. Violin plots for slice-wise diffusion-based model comparison on **(C1)** Test Set 1(N=583, 5 datasets) and **(C2)** Test Set 2 (N=27,719, 3 datasets). The violins represent kernel density estimates of the error distribution with a text overlay of mean values. Wider sections of the violins indicate a higher probability density at that error level. The diffusion 5-slice ensemble (AgeDiffuse-5) consistently performed with the highest accuracy among all models on both test sets (C1.1-C2.1). **C3.** Violin plots for prediction error distribution for each chronological age, divided by Test Set 1/Test Set 2. AgeDiffuse-5 demonstrated strong performance across the age range, with mild performance degradation for subjects older than 25 years (See Supplementary Figure S6 and Supplementary A4. Outlier Analysis).

Recent studies have proposed bias correction for deep learning regression models given the tendency for models to underestimate older age and overestimate younger age ^22^, albeit this correction strategy is controversial ^23^. We investigated brain age bias correction and found that it did not improve prediction accuracy (See Supplementary material A3. Age-Bias Correction).

### 2.2. Brain age and brain structure volumes

Interpretability of deep learning algorithms is clouded by the black-box nature of hidden layers^24^, and brain age models to-date have not investigated the underlying biological and anatomical bases of predictions. To improve the understanding of underlying factors contributing to brain age prediction, we analyzed associations with brain substructure volumes derived from Bethlehem et al ^25^ within overlapping patients from both studies for (N=25,096, age mean 12.2, Figure 3). We found that, graphically, chronologic age and predicted brain age had similar associations with brain substructure changes over development. We then examined how brain age gap, defined as per Eq (1), is associated with brain substructure volumes.

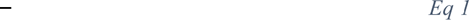

**Figure 3.**
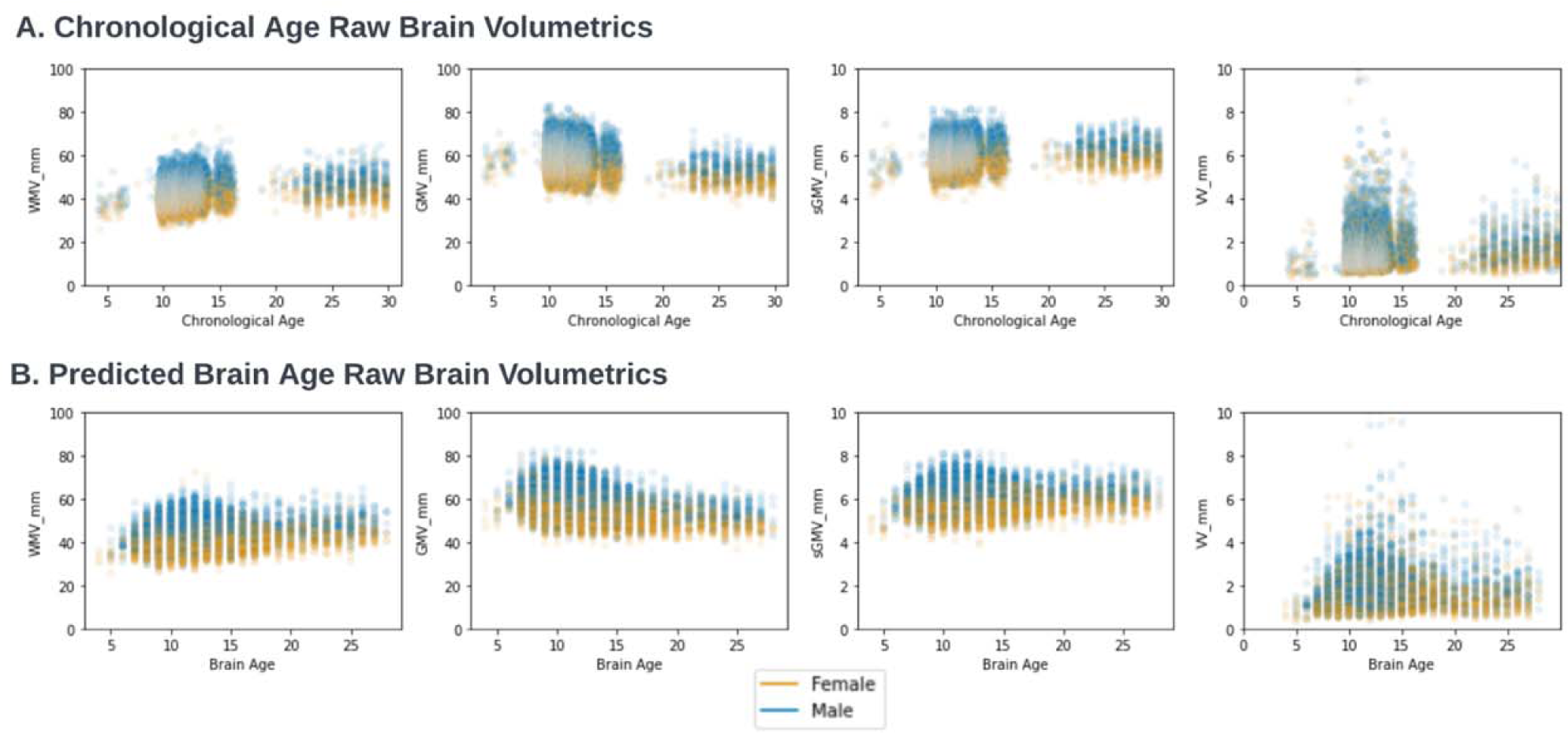
**A.** Deep Learning Brain Age and Structural Tissue Volumes. Brain structural tissue volumes for white matter (WMV), grey matter (GMV), total subcortical grey matter volume (sGMV), and ventricles (VV) are plotted for each cross-sectional control scan as a function of (**A**) chronological age and (**B**) predicted brain age using AgeDiffuse-5.

Specifically, we investigated whether ‘younger brain’ and ‘older brain’ outliers, defined as predicted brain age >1 standard deviation above or below the mean prediction for a given chronological age and sex, were associated with brain substructure volumes. We found that younger brain outliers had increased gray matter volume (GMV) and decreased white matter volume (WMV) and ventricle volume (VV), and older brain outliers had decreased subcortical gray matter volume (sGMV) and GMV, and increased VV (Mann-Whitney U test <0.003 for each, Figure 4A). Effect sizes were largest for VV and GMV for ‘older brain’ (Cohen’s *d*LJ>LJ0.2, Figure 4B).

**Figure 4.**
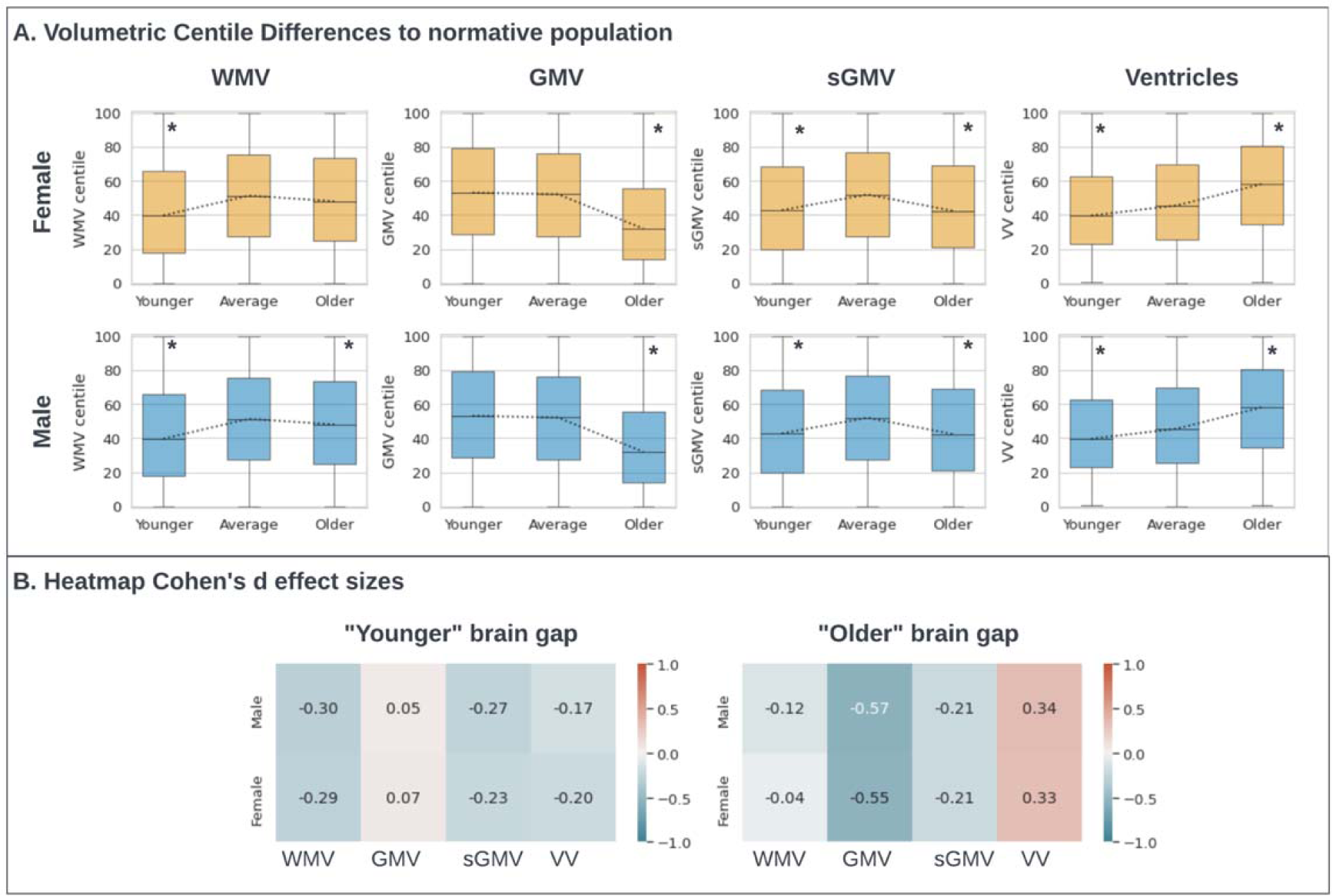
**A.** Box plots for brain age gap and association with brain substructure volumes. Brain age gap was defined as predicted brain age minus chronological age. “Younger” brain age gap was defined as predicted brain age >1 standard deviation below the mean; “Older” brain age gap was defined as predicted brain age >1 standard deviation above the mean. The “average” group was defined as those subjects whose brain age gap lies within one standard deviation. Pairwise tests for significance were based on the Mann-Whitney U-test, and P values were adjusted for multiple comparisons using the Bonferroni correction. Significant differences (with corrected PL<L0.003) are highlighted with an asterisk**. B.** Heatmap of Cohen’s d effect sizes comparing brain age outliers versus within normal range, stratified by gender and key volumetric measures from MRI. VV = cerebrospinal fluid, WMV = white matter volume, GMV = gray matter volume, sGMV = total subcortical grey matter volume.

To determine how brain substructure volume was comparatively associated with chronological versus brain age, we compared two multivariable linear regression models with brain substructure volumes and sex as independent variables and chronological age or predicted brain age as dependent variables. We found that brain substructure volume was more associated with brain age than chronological age (R-squared: 0.37 vs R-squared: 0.47; See Supplementary material A2. Linear model diagnostics).

### 2.3. Longitudinal brain age evaluation

A barrier to the clinical utility of brain age models is that, due to data availability, models are developed on cross-sectional data, yet the clinical impact would be strengthened by the ability to track individual brain age over time (and how exposures modify individual-level brain age). There is concern that brain age prediction derived from cross-sectional data does not generalize to individual-level brain age change ^17^. To investigate this, we applied AgeDiffuse-5 to longitudinal data available within the ABCD dataset, where each subject contains 3 MRI time points at roughly 2-year intervals. On longitudinal analysis, we found that predicted brain age tracked directionally with chronologic age, with a slight underestimation of chronological age that was within the margin of algorithm expected prediction error (Figure 5A).

**Figure 5.**
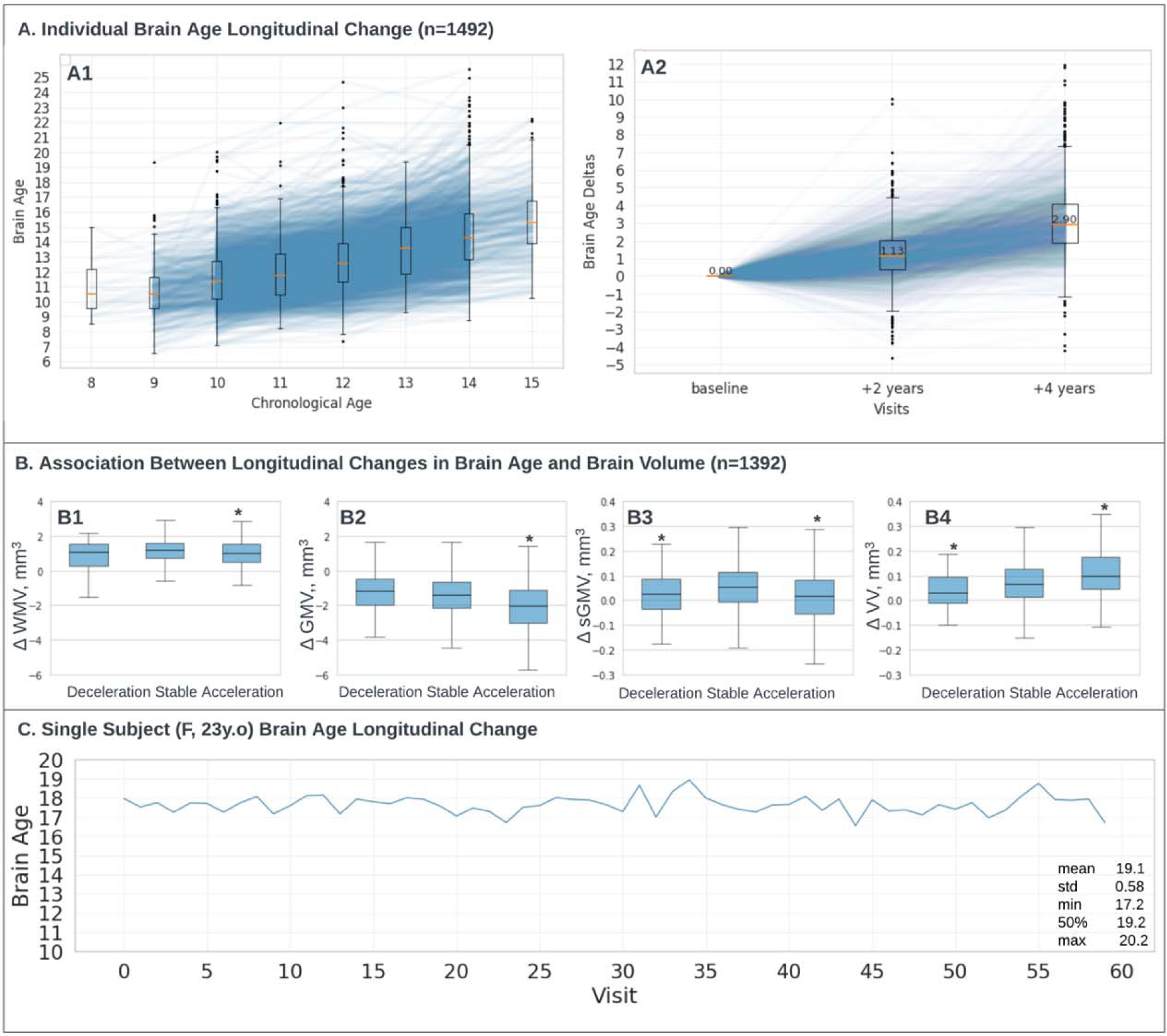
**A.** Individual Brain Age Longitudinal Change (n=1492). A1: Individual lines show brain age longitudinal change for 1492 subjects (ABCD dataset ^26^) who had 3 visits within 2 years in between with boxplot overlay. A2: Individual brain age changes in-between visits: baseline, +2 years timestamp was computed as the brain age difference for each subject between the second and baseline visit; +4 years timestamp was computed as the brain age difference for each subject between the third and baseline visit. B. Association Between Longitudinal Changes in Brain Age and Brain Volume (n=1392). The study examined the relationship between changes in brain age and changes in brain volume over time in 1,392 participants. Significant volumetric variables are marked with an asterisk (P values were adjusted for multiple comparisons, PL<L0.006, See Methodology “Longitudinal Brain Age Analysis”) **C.** The participant (Female, 23 years: 28andme dataset ^27^) underwent daily testing for two studies of 30 consecutive days with one year in between (60 scans in total). The mean predicted brain age is 19.1 years, with a standard deviation of 0.58. VV = cerebrospinal fluid, WMV = white matter volume, GMV = gray matter volume, sGMV = total subcortical grey matter volume.

We further examined the relationship between changes in brain age and brain substructure volumes over time in 1,392 subjects with available data. We found that the rate of change in brain age between subsequent MRI timepoints was associated with the rate of changes in brain substructure volumes over the same time interval (Figure 5B). Accelerated brain aging (i.e. change >1 standard deviation above the mean) was associated with an increased rate of growth in VV and a decreased rate of growth in sGMV, GMV, and WMV. Decelerated brain aging (i.e. change <1 standard deviation above the mean) was associated with a decreased rate of growth in sGMV and VV (adjusted P<0.006, Figure 5B, see Methods “Longitudinal Brain Age” section).

Finally, to demonstrate the stability of brain age predictions at the intra-patient level, we applied AgeDiffuse-5 for a single female participant tested over 60 days with daily MRI across two studies one year apart (28andme dataset ^27^). The mean predicted brain age was 19.1 years with a standard deviation of 0.58 across (**Error! Reference source not found.**C). The low standard deviation indicates consistent predictions across the 60 test days, with no observable trends in predicted age or error over time.

## 3. Discussion

Imaging-based brain age prediction in developing humans may have far-reaching clinical applications, though clinical translation has been limited by small datasets, unclear generalizability, and lack of reproducible models. In this study, we aggregated that largest to-date dataset of MRI scans for children through adulthood to develop and rigorously validate a diffusion-based regression neural network (AgeDiffuse) for brain age prediction. We found that AgeDiffuse, ensembled over multiple MRI slices among scans from a multi-institutional repository, demonstrated highly accurate and generalizable brain age prediction, outperforming current state-of-the-art models. AgeDiffuse was subject to two-tier validation across multiple datasets, and implementable code has been released open source as a resource for the scientific and clinical communities. Our results show that ensembling across axially sampled MRI slices can improve performance and that a technique where slice-based outlier predictions are excluded before averaging improves generalizability. Such a technique could enable accurate brain age prediction in patients with focal brain pathologies (e.g. tumors, vascular malformations, stroke), as the model would exclude slices with aberrant prediction. Additionally, we found that application of AgeDiffuse to longitudinal data was reliable and that the brain age prediction was driven, in part, by interpretable brain substructure volume changes that are associated with development. We believe this model is positioned for investigation in various pediatric conditions to track and predict brain development and neurocognitive outcomes in various diseases (e.g. brain tumors, endocrine dysfunction) and/or interventions (e.g. radiation therapy, hormonal therapies) that may affect normal development and neurocognitive outcomes.

Brain age tracking may reveal clinically relevant states, such as changes in the neurocognition ^5^, that could guide interventions and triage patients for escalated care. Previous studies have linked the brain age gap to various biomedical factors and lifestyle variables in healthy cohorts ^28–30^. Large-scale datasets have recently enabled the development of normative growth charts for key structural MRI metrics across ages, providing an essential reference for quantifying individual variation ^25^. These brain charts identify neurodevelopmental milestones, show reliability across scans, and can benchmark deviations in disorders. In this study, for the first time, we demonstrate that DL brain age prediction is associated with substructure volume changes that signify age-related atrophy at the individual-level. Our findings suggest that DL brain age and substructure volumetrics are likely complementary measures, though additional research should examine how much incremental information is added by DL brain age compared to structural volumetrics when predicting neurocognitive endpoints.

In the context of children and developing humans in the early part of the lifespan, several DL methods have emerged for age inference directly from 3D images, eliminating the need for prior feature extraction ^9^. Mendes et al ^10^ achieved an average 10-fold average Mean absolute error(MAE) of 1.57 years using 3D VGG16, utilizing data from two public datasets (ABIDE-II, N=580, and ADHD-200, N=922) covering an age range of 6 to 20 years. He et al ^12^ compared the performance of 2D- ResNet18+LSTM and 3D neural networks, reporting an MAE of 1.14 years versus 2.64 years on an external cohort with subjects aged 0 to 6 years (private dataset, N=428). Hong et al. ^13^ MAE of 67.6 days on an internal held-out test set of 44 subjects aged 0 to 5 years, utilizing a 3D CNN approach. Additionally, Hu et al. ^11^ proposed a 3D CNN model, demonstrating an average MAE of 1.01 years in a 5-fold cross-validation on 880 subjects (ABIDE I and II, ADHD200), spanning ages 6 to 18 years. However, only one of these methods has publicly available code with no model weights publicly available ^13^, and none have compared model generalization across multiple studies that were not included in the model training process. The focus on narrow age ranges and lack of rigorous evaluation on heterogeneous public datasets raises questions about model generalizability and reproducibility. While we were not able to directly benchmark AgeDiffuse to the models due to a lack of implementable code, we utilized three comparison approaches with similar, established 2D and 3D CNN architectures and optimized them with transfer and self-supervised learning. We found that diffusion-based model performance – even without ensembling – had improved performance. We hypothesize that the brain age correction procedure does not generalize well on unseen datasets and does not capture the non-linear, complex relationship between brain age and chronological age, unlike deep learning.

Our study highlights the challenges of brain age model generalization and has several important limitations. We noted that brain age prediction tends to become less precise in older age ranges, likely due to developmental and environmental heterogeneity ^31–33^. Specifically, we observed a performance drop in one of the smaller external validation datasets (WU1200), with an age range of 22 – 29 years. Notably, this population also had differences in substructure volumetrics, indicating that the performance drop may be due more to true population differences than problems with the model (Supplementary Material A4). These findings have been noted previously ^31–33^, and have implications for the utility of brain age in older populations. They also suggest that individual-level longitudinal trajectories of brain age may be more informative than snapshots compared to a general population. We were able to establish feasibility of longitudinal analysis within the ABCD cohort, although this was limited to age ranges 8 – 16, and further work is ongoing to evaluate longitudinal changes over longer intervals. Secondly, the aggregated MRI dataset might have a bias towards North American and European populations. This is a common pitfall of healthcare inequity that must be addressed by increasing the number of studies in other demographics. Moving forward, curating test sets that capture wide pediatric age ranges and those with real-world clinical data will better assess model performance for diverse real-world utilization, and we would recommend pilot testing in underrepresented patient groups prior to implementation ^34^. Additionally, utilizing multiple imaging modalities (for example, T1w and T2) could help to refine model prediction further.

## 4. Conclusions

In this work, we developed and rigorously validated an accurate brain age prediction model, AgeDiffuse, for children through adulthood using diffusion regression on multiple datasets. We demonstrated that this approach could be feasibly applied to longitudinal data to track individual brain age changes over time. Further analyses suggested that deep learning brain age and substructure volumetrics carry complementary information. With this study, we release, to our knowledge, the first fully implementable deep learning brain age algorithm to the scientific community. Independent validation of our model in the context of various conditions with longitudinal cohorts and clinical endpoints is needed to maximize the impact of deep learning-based brain age prediction for children through adulthood.

## 5. Materials and methods

### Dataset

We curated T1w MRIs without contrast enhancement from 16 datasets and stratified them by age so that each age had 100 scans per year maximum in the training set, to avoid data imbalance during the training (ABCD^35^, ABIDE ^36^, AOMIC^37^, Baby Connectome^38^, Calgary^39^, ICBM^40^, IXI^41^, NIMH^42^, PING^43^, Pixar^44^, SALD^45^, NYU2(CoRR) ^46^, Healthy Adults^47^; Long579^48^, WU1200^49^; see Supplementary Material A5). To create robust train and test sets, we divided the data into training, validation, and test sets using a rough 70/15/15 split. When splitting the data, we matched the age distribution coverage between the training and test sets as closely as possible. This ensured that both sets had similar representation across the full range of ages. At the same time, we preserved the integrity of each original dataset by keeping all subjects from a given source together in either the training or test set. This avoided contaminating the test data with subjects from datasets used in training. The training data consisted of 8 datasets totaling 4,549 subjects (Figure 2, Panel A2). We held out 5 separate datasets with 583 total subjects as our first test set (Figure 2, Panel A3). We also created a larger second test set using 3 primary datasets with 27,719 subjects (Figure 2, Panel A4).

### Image Preprocessing and Registration

Scans were co-registered to MRI age-dependent T1-weighted asymmetric brain atlases, generated from the NIH-funded MRI Study of Normal Brain Development (hereafter, NIHPD, for NIH pediatric database ^50^) with rigid registration using SlicerElastix^51^ (Elastix generic rigid preset). All MRIs were skull-stripped using HD-BET^52^. MRI images were rescaled to 1-mm isotropic voxel size to preserve anatomical size differences using the itk-elastix Python package ^53^. N4 bias field correction was performed using the simple-itk Python library. We then normalized MRI images, performed median filtering, removed background pixels using Otsu filtering, and standardized the intensity scale. After preprocessing, we identified axial slices with at least 1% non-zero voxels to ensure consistent anatomical coverage across subjects. We extracted five equidistant percentile slices from these valid slices along the inferior-superior axis - the 25th, 37.5th, 50th, 62.5th, and 75th percentiles. The 50th percentile median slice focused on central structures, while lower and higher percentile slices sampled inferior and superior regions. This multi-slice approach provided an anatomically distributed sampling of the pediatric brain for 2D deep learning analyses.

### Regression Dual-Guidance Diffusion Model

The overall pipeline is shown in **Error! Reference source not found.**B. We modified the dual-guidance diffusion model architecture for medical image classification(DiffMIC) proposed by Yijun Yang et al^19^ into a regression task by changing the loss function to mean squared error and adding a final fully connected layer. Additionally, we added an early stopping rule with patience=50 for both models. We trained all models separately on an A6000 Nvidia GPU; further technical details and code can be found on the GitHub repository that would be made public upon acceptance.

### Model Ensembling

We conducted experiments comparing simple model averaging with varying ensemble sizes and outlier exclusion to evaluate different ensembling techniques for improving predictive uncertainty. Ensembles of sizes 3 and 5 were constructed by training identical model architectures for different slice quantiles. We investigated an “outlier exclusion” ensembling technique to mitigate the effect of outlier scans on age prediction. We hypothesized that these outliers were likely due to image artifacts, poor quality scans, MRI registration or other out-of-distribution characteristics. For the outlier exclusion ensemble, 5 models were trained, and each model produced a brain age prediction for a given input. The standard deviation of the predictions from the 5 models was calculated. Any individual model prediction that was an outlier meaning it deviated markedly from the ensemble average, was excluded. The remaining model predictions were averaged to produce the final consensus prediction. All ensembles were evaluated by two-tiered external validation (Figure 2).

### Brain Substructure Volumetrics

We used the centile definition described in Bethlehem et al ^25^; for details on the normative growth charts, please refer to the original publication. We obtained a total of 25.097 overlapping scans from datasets ABCD^35^, IXI^41^, Pixar^44^, SALD^45^, WU1200^49^; see Supplementary Material A5). Four key volumetric centile measurements (WMV, GMV, sGMV, VV) were compared pairwise between “older”/”younger” and “average” age groups for each gender. Brain age gap was defined as predicted brain age minus chronological age. “Younger” brain age gap was defined as predicted brain age >1 standard deviation below the mean; “Older” brain age gap was defined as predicted brain age >1 standard deviation above the mean. The “average” group was defined as those subjects whose brain age gap lies within one standard deviation. Pairwise Mann-Whitney U tests were used to compare the older group to the average age group for each volumetric and gender. Bonferroni correction was applied to adjust for multiple comparisons (adjusted alpha = 0.05/16 = 0.003125). Cohen’s d effect sizes were calculated to quantify the standardized mean difference between groups for each volumetric and gender.

### Longitudinal Brain Age Analysis

To calculate the association between longitudinal changes in brain age and brain volume over time in 1,392 participants, we calculated the rate of volumetric measures change (WMV, GMV, sGMV, VV) for each time point and each subject and calculated their brain age using AgeDiffuse-5. The acceleration values were then categorized as “Accelerated”, “Decelerated”, or “Stable” based on standard deviation thresholds. For each volumetric, pairwise two-sided Mann-Whitney U tests compared the “Stable” group to “Accelerated”/ “Decelerated”. Bonferroni correction was applied to adjust for multiple comparisons across the 4 volumetrics (PlJ<lJ0.006)

### Performance Evaluation and Statistical Analysis

The primary endpoint was the mean average absolute error of predicted age compared to chronological age (ground truth). Violin and box plots with median errors were used for visual comparison. Associations between substructures and brain age or chronological age were evaluated with multivariable logistic regression. Model goodness of fit was evaluated by comparing R^2^ values (See Supplementary Material A2). Pairwise tests for significance were based on the two-sided Mann-Whitney U-test, and P values were adjusted for multiple comparisons using the Bonferroni correction.

## Abbreviations

DL: Deep learning
MRI: Magnetic resonance imaging
MAE: Mean absolute error
IQR: Interquartile range 95%
CI: 95% confidence interval
SoTa: state-of-the-art
DiffMIC: dual-guidance diffusion model for medical image classification
AgeDiffuse: Novel regression dual-guidance diffusion model for brain age prediction
VV: cerebrospinal fluid
WMV: white matter volume
GMV: gray matter volume
sGMV: total subcortical grey matter volume
CNN: convolutional neural network

## Data availability

The complete dataset (Supplementary Material A5) aggregated for this study contains primary datasets that differ widely in terms of their “openness,” i.e., their availability for secondary use without restrictions or special efforts by the primary study team. Preliminary studies ranged from fully open and downloadable datasets in the public domain to more restricted datasets that could only be used for specific purposes, under separate agreements, or after special efforts had been made to provide data in shareable form.

## Code availability

The model training and testing code will be made available in the study git repository upon acceptance.

## Ethical Approval and Informed Consent

The datasets were anonymized and not collected by the investigators, in which case the work is classified as non-human research.

## Authors’ Contributions

Conceptualization and Study Design: AZ, BHK

Data collection/curation: AZ, BHK, RBC, SV, JS, RAIB

Investigation: AZ, BHK

Code, Software: AZ

Methodology, Formal Analysis, Visualizations (Figures): AZ, BK

Data Interpretation: AZ, BHK

Manuscript Writing - original draft: AZ, BHK

Manuscript Writing - review & editing: AZ, DT, ZY, KXL, JL, SV, RBC, JS, RAIB, RHM, SM, DAHK, TYP, HJWLA, BHK

Project administration: BHK, HJWLA

Resources: BHK, HJWLA, TYP

Supervision: BHK, HJWLA

All authors have substantively revised the work, reviewed the manuscript, approved the submitted version, and agreed to be personally accountable for their contributions.

## Acknowledgments

**Conflict of interest/Competing interests.** JS and RAIB hold equity in and serve on the board of Centile Bioscience. All other authors declare no conflict of interest.

**Consent for publication.** All authors consent for publication.

## Supplementary Material

### A1. Model hyperparameter tuning

#### RagImageNet Finetuning

The RadImageNet database is an open-access medical imaging database. It was designed to improve transfer learning performance on downstream medical imaging applications^20^. We used RadImageNet pretrained ResNet50 backbone and added 3 fully connected layers (sizes: 1024, 128, 1) in combination with dropout layers (0.5) and fine-tuned unfreezing all layers using Adam optimizer learning rate 1e-3 that reduces on plateau and MAE loss, with an early stopping rule (patience=10) and batch size 32.

#### ModelGenesis Finetuning

We pre-trained ModelGenesis 3D U-net backbone in a self-supervised way on the brain MRI scans as described in ^21^. We further used the encoder with one fully connected layer (size:512) for finetuning using SGD optimizer with learning rate 1e-5 that reduces on plateau for 20 epochs and MSE loss. We used batch size 1 and downscaled MRI T1w to [64,64,64] patch size with resolution [2,2,2].

**Figure S6.**
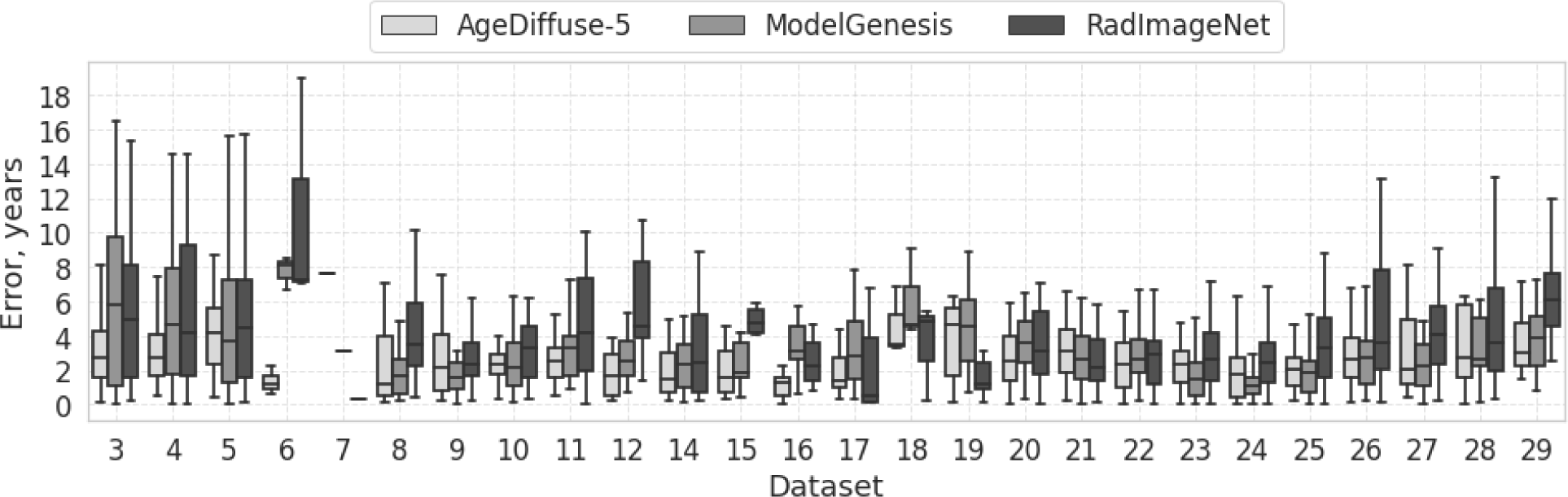
Box plots for prediction error distribution of different models for each chronological age on Test Set 1(n=583) AgeDiffuse-5 demonstrated strong performance across the age range, with mild performance degradation for subjects older than 25 years.

### A2. Linear model diagnostics

We analyzed linear model fit using the statsmodels python package to identify potential problems that can occur from fitting linear regression model to non-linear relation. We compared two linear regressions with VV, WMV, sGMV, GMV and sex variables as predictors and chronological age versus predicted brain age as dependent variables and found that the brain age variable had a higher R-squared value (R2:0.37, F-stat:2936, AIC:1.1e+5, Figure S7 vs R2:0.48, F-stat:4587, AIC: 1.1e+5, Figure S8), indicating a stronger correlation between structural changes and predicted brain age as compared to chronological age.

**Figure S7.**
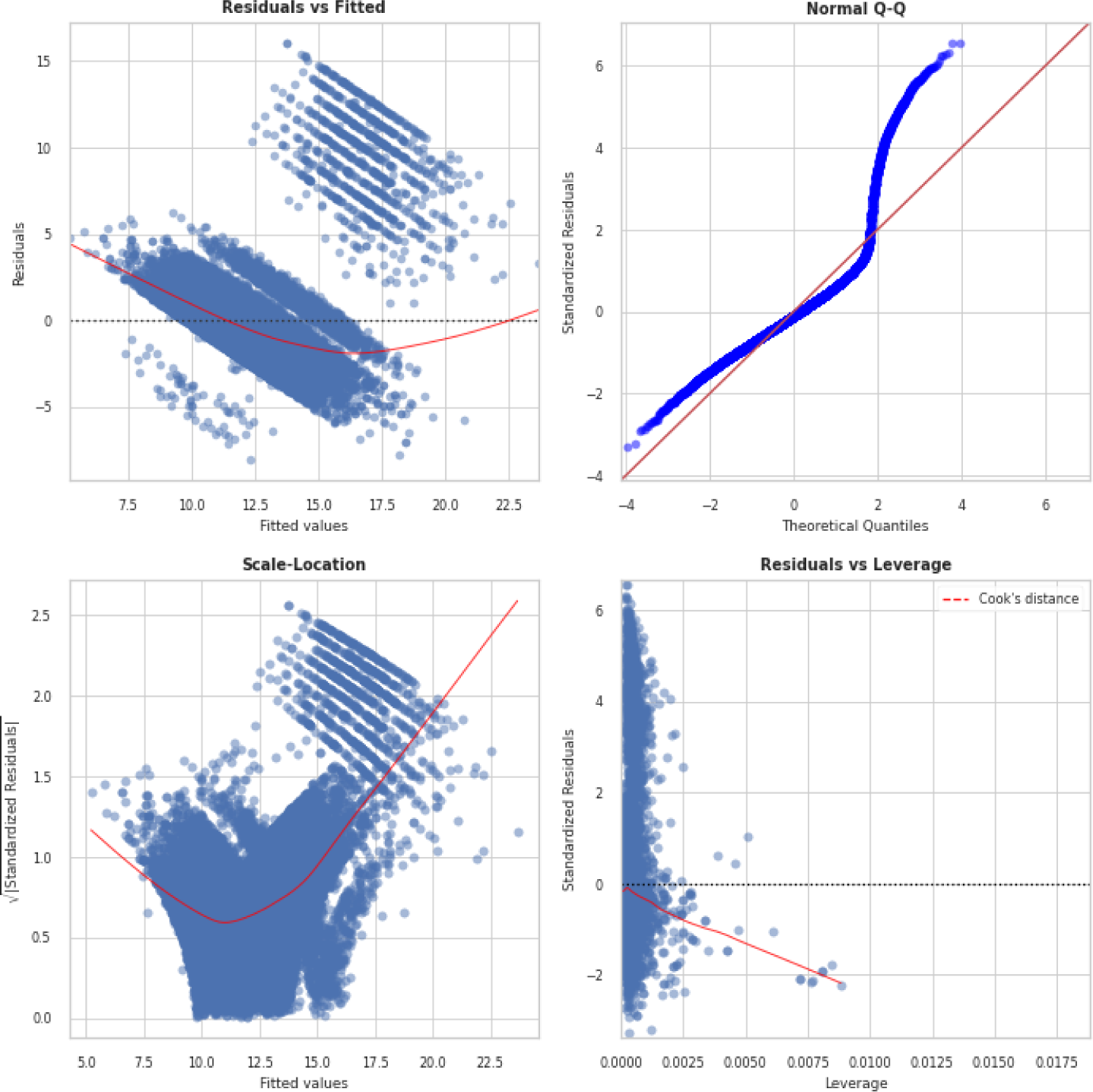
Chronological Age Linear Model Diagnostics(R2:0.37, F-stat:2936, AIC:1.1e+5). **Top left:** Residual vs Fitted values. In the graph, a red (roughly) horizontal line would be an indicator that the residual has a linear pattern. **Top right:** Standardized Residual vs Theoretical Quantile to check if residuals are normally distributed visually. **Bottom left:** Sqrt(Standardized Residual) vs Fitted values to check homoscedasticity of the residuals, with non-horizontal scatter suggesting the variance of errors is not constant. **Bottom right:** Residual vs Leverage Points falling outside the Cook’s distance curves are considered observations that can sway the fit.

**Figure S8.**
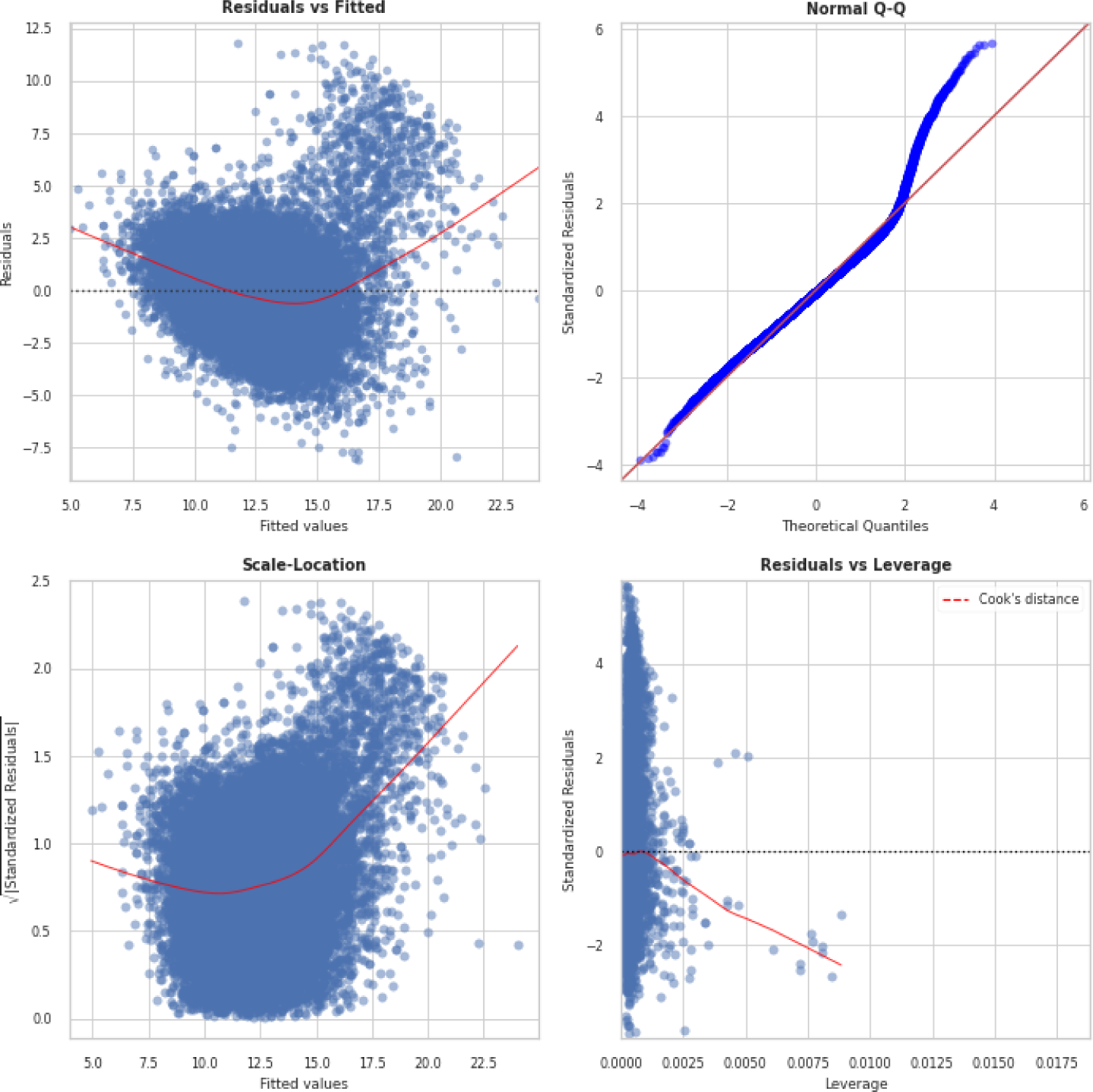
Predicted Brain Age Linear Model Diagnostics(R2:0.48, F-stat:4587, AIC: 1.1e+5). **Top left:** Residual vs Fitted values. In the graph, a red (roughly) horizontal line would be an indicator that the residual has a linear pattern. **Top right:** Standardized Residual vs Theoretical Quantile to check if residuals are normally distributed visually. **Bottom left:** Sqrt(Standardized Residual) vs Fitted values to check homoscedasticity of the residuals, with non-horizontal scatter suggesting the variance of errors is not constant. **Bottom right:** Residual vs Leverage Points falling outside the Cook’s distance curves are considered observations that can sway the fit.

### A3. Age-Bias Correction

We used the linear bias correction method described by Smith et al. ^54^ for bias correction for the gap. Such a bias correction is valuable for most brain-age prediction studies, as there is normally an underfitting of the prediction due to problems such as regression dilution and non-Gaussian age distribution. Defining to be chronological age and the predicted age, we fitted a linear regression = + to the left-out validation set (with labels). The corrected predicted age is estimated by =(−)/ This method requires (at the point of estimating a and b from x and y) that the chronological ages are known. For the two external test sets, we assumed that and are generalizable. We used the coefficients (a=1.1, b=-2.2) fitted on Test Set 1 to estimate the corrected brain-age gap (Figure S9). We found that brain age correction does not improve MAE on Test Set 2 (no correction MAE = 1.9 years; with correction MAE = 2.6 years). We hypothesize that the brain age correction procedure does not generalize well on unseen datasets and does not capture the non-linear, complex relationship between brain age and chronological age, unlike deep learning, and therefore we used “raw” brain age predictions for all the analysis in this paper.

**Figure S9.**
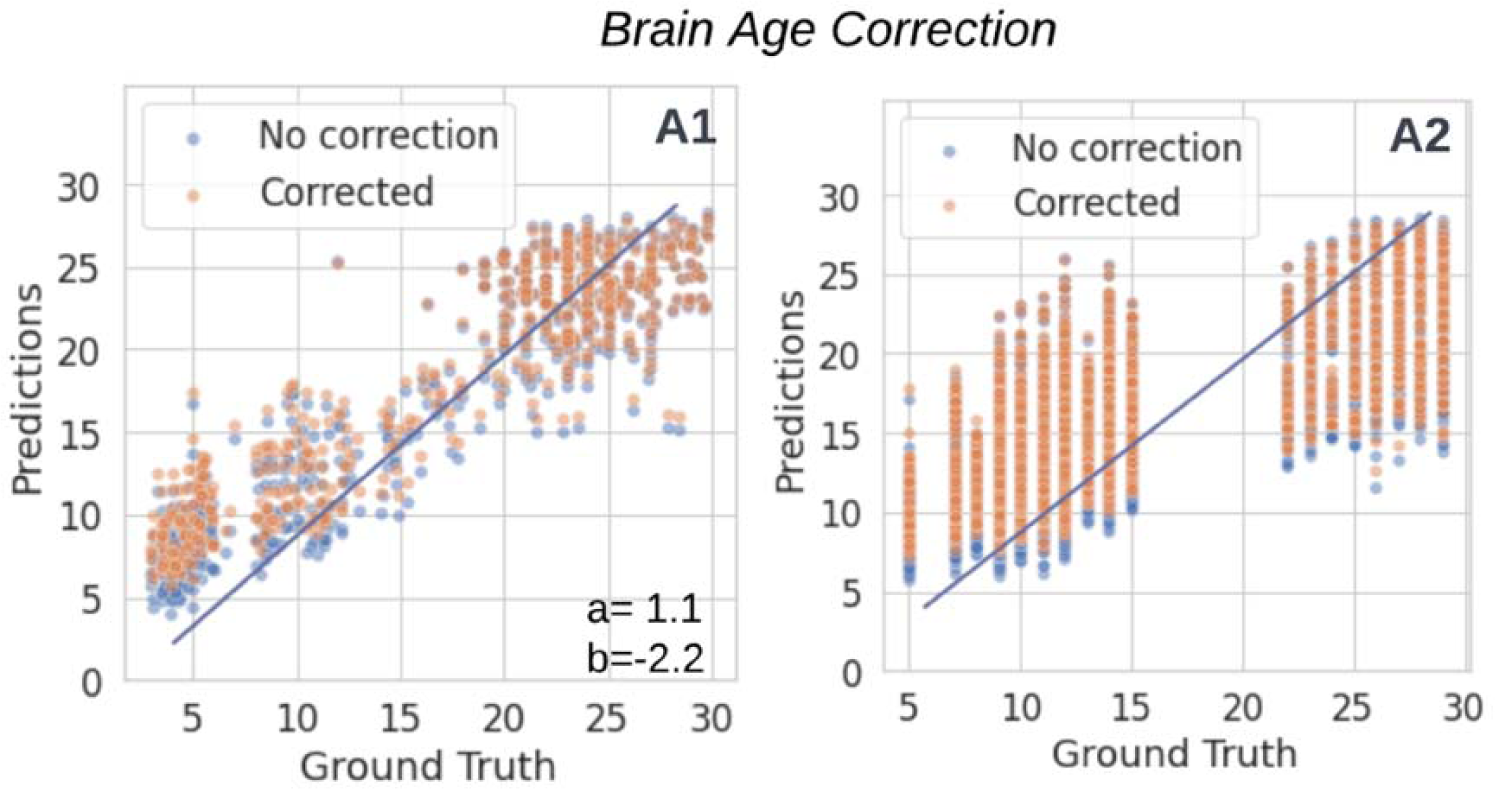
Brain age-correction scatterplots with fitted regression line. Panel A1: Test Set 1(N=583), Panel A2: Test Set 2 (N=27.719). We fitted linear regression using Test Set1, a=1.1, b=-2.2.

### A4. Outlier Analysis

To investigate performance drop in the WU1200 dataset, we compared key volumetric measures in mm^3^(VV, WMV, sGMV, GMV) between WU1200 (N=620) and an adult test set (age 22+, N=226; datasets: IXI^41^, Pixar^44^, SALD^45^). We used pairwise Mann-Whitney-U test and calculated the adjusted alpha to account for multiple comparisons using Bonferroni correction (adjusted P=0.0125). We also calculate Cohen’s d effect size to quantify the standardized mean difference. We found that VV (Cohen’s d=0.98), sGMV (Cohen’s d=0.25) were significantly higher and WMV (Cohen’s d=-0.49) was significantly lower in WU1200. This could indicate developmental differences, highlighting that the observed performance drop may be due more to true population differences than problems with the model.

### A5. Primary Datasets

**Table S2.**
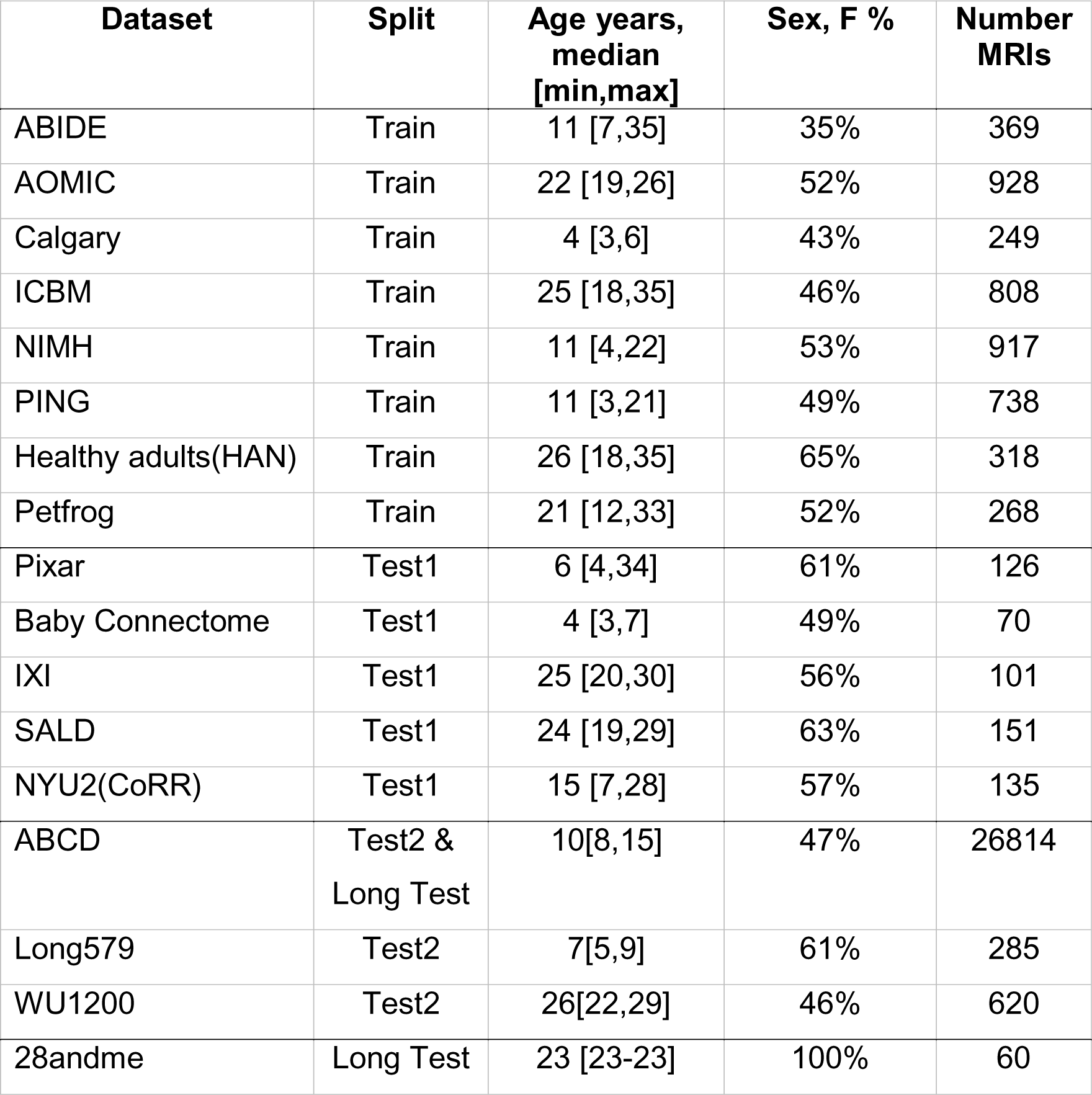
Dataset demographics.

| **Dataset** | **Split** | **Age years,**<br>**median**<br>**[min,max]** | **Sex, F %** | **Number**<br>**MRIs** |
| --- | --- | --- | --- | --- |
| ABIDE | Train | 11 [7,35] | 35% | 369 |
| AOMIC | Train | 22 [19,26] | 52% | 928 |
| Calgary | Train | 4 [3,6] | 43% | 249 |
| ICBM | Train | 25 [18,35] | 46% | 808 |
| NIMH | Train | 11 [4,22] | 53% | 917 |
| PING | Train | 11 [3,21] | 49% | 738 |
| Healthy adults(HAN) | Train | 26 [18,35] | 65% | 318 |
| Petfrog | Train | 21 [12,33] | 52% | 268 |
| Pixar | Test1 | 6 [4,34] | 61% | 126 |
| Baby Connectome | Test1 | 4 [3,7] | 49% | 70 |
| IXI | Test1 | 25 [20,30] | 56% | 101 |
| SALD | Test1 | 24 [19,29] | 63% | 151 |
| NYU2(CoRR) | Test1 | 15 [7,28] | 57% | 135 |
| ABCD | Test2 &<br>Long Test | 10[8,15] | 47% | 26814 |
| Long579 | Test2 | 7[5,9] | 61% | 285 |
| WU1200 | Test2 | 26[22,29] | 46% | 620 |
| 28andme | Long Test | 23 [23-23] | 100% | 60 |

### ABCD

Data used in the preparation of this article were obtained from the Adolescent Brain Cognitive Development SM (ABCD) Study (https://abcdstudy.org), held in the NIMH Data Archive (NDA). This is a multisite, longitudinal study designed to recruit more than 10,000 children age 9-10 and follow them over 10 years into early adulthood. The ABCD Study® is supported by the National Institutes of Health and additional federal partners under award numbers U01DA041048, U01DA050989, U01DA051016, U01DA041022, U01DA051018, U01DA051037, U01DA050987, U01DA041174, U01DA041106, U01DA041117, U01DA041028, U01DA041134, U01DA050988, U01DA051039, U01DA041156, U01DA041025, U01DA041120, U01DA051038, U01DA041148, U01DA041093, U01DA041089, U24DA041123, U24DA041147. A full list of supporters is available at https://abcdstudy.org/federal-partners.html. A listing of participating sites and a complete listing of the study investigators can be found at https://abcdstudy.org/consortium_members/. ABCD consortium investigators designed and implemented the study and/or provided data but did not necessarily participate in the analysis or writing of this report. This manuscript reflects the authors’ views and may not reflect the opinions or views of the NIH or ABCD consortium investigators. The ABCD data repository grows and changes over time. The ABCD data used in this report came from the fast-track data release. The raw data are available at https://nda.nih.gov/edit_collection.html?id=2573. Instructions on how to create an NDA study are available at https://nda.nih.gov/training/modules/study.html). Additional support for this work was made possible from supplements to U24DA041123 and U24DA041147, the National Science Foundation (NSF 2028680), and Children and Screens: Institute of Digital Media and Child Development Inc. ^35^

### ABIDE

ABIDE II involves 19 sites, ten charter institutions and seven new members, overall donating 1114 datasets from 521 individuals with ASD and 593 controls (age range: 5-64 years). These data were openly released to the scientific community on June 2016. In accordance with HIPAA guidelines and 1000 Functional Connectomes Project / INDI protocols, all datasets are anonymous, with no protected health information included. Consistent with its popularity in the imaging community and prior usage in FCP/INDI efforts, the NIFTI format was selected to store the ABIDE II MRI datasets (http://fcon_1000.projects.nitrc.org/indi/abide/abide_II.html). With the exception of a single collection (IP1, 1.5 Tesla), all MRI data were acquired using 3 Tesla scanners^36^.

#### Aomic

The Amsterdam Open MRI Collection (AOMIC, https://openneuro.org/datasets/ds003097/versions/1.2.1) is a collection of three datasets with multimodal (3T) MRI data, including structural (T1-weighted), diffusion-weighted, and (resting-state and task-based) functional BOLD MRI data, as well as detailed demographics and psychometric variables from a large set of healthy participants (N = 928, N = 226, and N = 216). Data from all three datasets were acquired on the same Philips 3T scanner (Philips, Best, the Netherlands) but underwent several upgrades in between the three studies ^37^.

#### Baby Connectome

The Baby Connectome Project (BCP: https://nda.nih.gov/edit_collection.html?id=2848) is a four-year study of children from birth through five years of age, intended to provide a better understanding of how the brain develops from infancy through early childhood and the factors that contribute to healthy brain development. This project is a research initiative of the Neuroscience Blueprint – a cooperative effort among the 15 NIH Institutes, Centers, and Offices that support neuroscience research. The BCP is supported by Wyeth Nutrition through a donation to the FNIH. Images are acquired on 3T Siemens Prisma MRI scanners using a Siemens 32-channel head coil at the Center for Magnetic Resonance Research (CMRR) at the University of Minnesota and the Biomedical Research Imaging Center (BRIC) at the University of North Carolina at Chapel Hill ^38^.

#### Calgary

The Preschool MRI study in The Developmental Neuroimaging Lab at the University of Calgary uses different magnetic resonance imaging (MRI) techniques to study brain structure and function in early childhood (https://osf.io/axz5r/files/osfstorage). All imaging for this dataset was conducted using the same General Electric 3T MR750w system and 32-channel head coil (GE, Waukesha, WI) at the Alberta Children’s Hospital in Calgary, Canada. Children were scanned either while awake and watching a movie, or while sleeping without sedation. The University of Cal- gary Conjoint Health Research Ethics Board (CHREB) approved this study (REB13-0020). T1-weighted images were acquired using an FSPGR BRAVO sequence with TR = 8.23 ms, TE = 3.76 ms, TI = 540 ms, flip angle=12 degrees, voxel size = 0.9=0.9=0.9 mm3, 210 slices, matrix size=512=512, field of view=23.0 cm. ASL images were acquired with the vendor supplied pseudo continuous 3D ASL sequence with TR = 4.56 s, TE = 10.7 ms, in-plane reso- lution of 3.5=3.5 mm2, post label delay of 1.5 s, and thirty 4.0 mm thick slices. The sequence scan time was 4.4 minutes ^39^

### ICBM

Data used in the preparation of this work were obtained from the International Consortium for Brain Mapping (ICBM) database (www.loni.usc.edu/ICBM). The ICBM project (Principal Investigator John Mazziotta, M.D., University of California, Los Angeles) is supported by the National Institute of Biomedical Imaging and BioEngineering. ICBM is the result of efforts of co-investigators from UCLA, Montreal Neurologic Institute, University of Texas at San Antonio, and the Institute of Medicine, Juelich/Heinrich Heine University - Germany. Data collection and sharing for this project was provided by the International Consortium for Brain Mapping (ICBM; Principal Investigator: John Mazziotta, MD, PhD). ICBM funding was provided by the National Institute of Biomedical Imaging and BioEngineering. ICBM data are disseminated by the Laboratory of Neuro Imaging at the University of Southern California ^40^.

### IXI

The data has been collected at three different hospitals in London:Hammersmith Hospital using a Philips 3T system (details of scanner parameters: http://brain-development.org/scanner-philips-medical-systems-intera-3t/), Guy’s Hospital using a Philips 1.5T system (details of scanner parameters: http://brain-development.org/scanner-philips-medical-systems-gyroscan-intera-1-5t/), Institute of Psychiatry using a GE 1.5T system (details of the scan parameters not available at the moment). The Thames Valley MREC granted ethical approval. The T1 and T2 images were acquired prior to diffusion-weighted imaging using 3D MRPRAGE and dual-echo weighted imaging ^41^.

### NIMH

The data used in this work was collected from the 5.1 release (https://nda.nih.gov/edit_collection.html?id=1151) . MRI scans were acquired using either General Electric or Siemens 1.5 Tesla scanners involving six sites or Pediatric Study Centers (PSC) in the United States. The Institutional Review Board at the University of Wisconsin-Madison also approved the analysis of the data of this human subject. Sequence type: 3D FLASH/SPGR; GE sequence: pulse sequence=SPGR, mode=3D; TR: 22 ms; TE: 10-11 ms; excitation pulse angle: 30 degrees; orientation: sagittal; FoV: 250mmISx250mmAP; matrix: 256 x 256 (x 124 - 180 slices); slices: 160- 180 slices of 1-1.5 mm thickness (cover entire head). Note that on GE systems with a 124-slice limitation, slice thickness should be adjusted to cover the entire head with 124 slices: signal averages: 1; scan time: 11.6 – 16.8 min ^42^.

### PING

The PING Data Resource(https://nda.nih.gov/edit_collection.html?id=2607) is the product of a multi-site project involving developmental researchers across the United States, including UC San Diego, the University of Hawaii UC Los Angeles Children’s Hospital of Los Angeles of the University of Southern California UC Davis Kennedy Krieger Institute of Johns Hopkins University Sackler Institute of Cornell University University of Massachusetts Massachusetts General Hospital at Harvard University and Yale University. The Data Resource includes neurodevelopmental histories, information about developing mental and emotional functions, multimodal brain imaging data, and genotypes for well over 1000 children and adolescents between the ages of 3 and 20. The PING imaging protocol takes advantage of key technologies developed for the consortium and builds on earlier methods development performed as part of the Biomedical Informatics Research Network (BIRN ^55^ and the Alzheimer’s Disease Neuroimaging Initiative (ADNI ^56^). Specifically, a standard PING scan session included: 1) a 3D T1-weighted inversion prepared RF-spoiled gradient echo scan using prospective motion correction (PROMO), for cortical and subcortical segmentation; 2) a 3D T2-weighted variable flip angle fast spin echo scan, also using PROMO, for detection and quantification of white matter lesions and segmentation of VV; 3) a high angular resolution diffusion imaging (HARDI) scan, with integrated B0 distortion correction (DISCO), for segmentation of white matter tracts and measurement of diffusion parameters; and 4) a resting state blood oxygenation level-dependent (BOLD) fMRI scan, with integrated distortion correction. Pulse sequence parameters used across (3 T) scanner manufacturers (GE, Siemens, and Phillips) and models were optimized for equivalence in contrast properties and consistency in image-derived quantitative measures ^43^.

#### Pixar

One hundred twenty-two 3.5–12-year-old children (M(s.d.)L=L6.7(2.3); 64 females) participated in the study (https://openfmri.org/dataset/ds000228/). Child and adult participants were recruited from the local community. All adult participants gave written consent; parent/guardian consent and child assent was received for all child participants. Recruitment and experiment protocols were approved by the Committee on the Use of Humans as Experimental Subjects (COUHES) at the Massachusetts Institute of Technology. Whole-brain structural and functional MRI data were acquired on a 3- Tesla Siemens Tim Trio scanner located at the Athinoula A. Martinos Imaging Center at MIT. Children under age 5 years used one of two custom 32-channel phased-array head coils made for younger (nL=L3, M(s.d.)L=L3.91(.42) years) or older (nL=L28, M(s.d.)L=L4.07(.42) years) children; all other participants used the standard Siemens 32-channel head coil. T1-weighted structural images were collected in 176 interleaved sagittal slices with 1 mm isotropic voxels (GRAPPA parallel imaging, acceleration factor of 3; adult coil: FOV: 256 mm; kid coils: FOV: 192 mm). Functional data were collected with a gradient-echo EPI sequence sensitive to Blood Oxygen Level Dependent (BOLD) contrast in 32 interleaved near-axial slices aligned with the anterior/posterior commissure and covering the whole brain (EPI factor: 64; TR: 2 s, TE: 30 ms, flip angle: 90°). This data was obtained from the OpenfMRI database, accession number is ds000228. Dataset version 1.0.2 ^44^

### SALD

The data was generated in the Southwest University Adult Lifespan Dataset (SALD), which comprises a large cross-sectional sample (n = 494; age range = 19-80) undergoing a multi-modal (sMRI, rs-fMRI, and behavioral). All data were collected at the Southwest University Center for Brain Imaging using a 3.0-T Siemens Trio MRI scanner (Siemens Medical, Erlangen, Ger- many). A magnetization-prepared rapid gradient echo (MPRAGE) sequence was used to acquire high-resolution T1-weighted anatomical images (repetition time=1,900ms, echo time=2.52ms, inversion time=900ms, flip angle=90 degrees, resolution matrix=256×256, slices=176, thickness =1.0mm, and voxel size=111mm3) ^45^.

### NYU2(CoRR)

The Consortium for Reliability and Reproducibility (CoRR, http://fcon_1000.projects.nitrc.org/fcpClassic/FcpTable.html) has aggregated 1,629 typical individuals’ resting state fMRI (rfMRI) data (5,093 rfMRI scans) from 18 international sites and is openly sharing them via the International Data-sharing Neuroimaging Initiative (INDI). In this study, we used a subset from CoRR study ”NYU 2” created by New York University (Di Martino, Kelly)^57^.

#### Healthy adults

The dataset was collected and shared under the NIMH Healthy Research Volunteer (RV) Study (Recruitment and Characterization of Healthy Research Volunteer for NIMH Intramural Studies NCT033046, https://openneuro.org/datasets/ds004215/versions/1.0.1). Data collection is ongoing, while data from 1,090 participants (155 with MRI) is shared. The MR protocol used was initially based on the ADNI-3 basic protocol, but was later modified to include portions of the ABCD protocol. Because there may be small changes in parameters from the standard ABCD/ADNI3 sequences, detailed sequence descriptions are shared in the BIDS source data directory. ^47^.

#### 28andMe

In this set of dense-sampling, deep phenotyping studies, we determined whether day-to-day variation in sex hormone concentrations impacts large-scale brain network connectivity. In Study 1 (sessions 1-30, 2018), the female participant was naturally cycling; in Study 2 (sessions 31-60, 2019), the participant was placed on an oral hormonal contraceptive regimen. The participant underwent a daily magnetic resonance imaging scan on a Siemens 3T Prisma scanner equipped with a 64-channel phased-array head coil. First, high-resolution anatomical scans were acquired using a T1- weighted magnetization prepared rapid gradient echo (MPRAGE) sequence (TR = 2500 ms, TE = 2.31 ms, TI = 934 ms, flip angle = 7°, 0.8 mm thickness) followed by a gradient echo fieldmap (TR = 758 ms; TE1 = 4.92 ms; TE2 = 7.38 ms; flip angle = 60°). Next, the participant completed a 10-minute resting-state fMRI scan using a T2*- weighted multi-band echo-planar imaging (EPI) sequence sensitive to the blood oxygenation level-dependent (BOLD) contrast (72 oblique slices, TR = 720 ms, TE = 37 ms, voxel size = 2 mm3, flip angle = 56°, multiband factor = 8). High-resolution anatomical scans were acquired using a T1-weighted magnetization prepared rapid gradient echo (MPRAGE) sequence (TR = 2500 ms, TE = 2.31 ms, TI = 934 ms, flip angle = 7°, 0.8 mm thickness) followed by a gradient echo fieldmap (TR = 758 ms; TE1 = 4.92 ms; TE2 = 7.38 ms; flip angle = 60°). A T2- weighted turbo spin echo (TSE) scan was also acquired with an oblique coronal orientation positioned orthogonally to the main axis of the hippocampus (TR/TE= 8100/50 ms, flip angle = 122°, 0.4 × 0.4 mm2 in plane resolution, 2 mm slice thickness, 31 interleaved slices with no gap, total acquisition time = 4:21 min) ^27^.

#### Long579

The public neuroimaging and behavioral dataset entitled “A longitudinal neuroimaging dataset on language processing in children ages 5, 7, and 9 years old” available on the OpenNeuro project (https://openneuro.org) and organized in compliance with the Brain Imaging Data Structure (BIDS). It includes 322 participants, recruited from the Austin, Texas. All neuroimaging data were collected using a Siemens Skyra 3LT MRI scanner located at The University of Texas at Austin Imaging Research Center. All images were acquired using a 64-channel head coil. Participants were positioned supine in the MRI scanner and foam pads were placed around the head to minimize movement. T1- weighted Magnetization Prepared - RApid Gradient Echo (MPRAGE) images were collected using GRAPPA, a parallel imaging technique based on k-space, and the following parameters: GRAPPA accel.factor PEL=L2, TRL=L1900 ms, TEL=L2.43Lms, field of viewL=L256Lmm, matrix sizeL=L256L×L256, bandwidthL=L180LHz/Px, slice thicknessL=L1Lmm, number of slicesL=L192, voxel sizeL=L1Lmm isotropic, flip angleL=L9°. ^58^

### WU1200

This HCP data release includes high-resolution 3T MR scans from young healthy adult twins and non-twin siblings (ages 22-35) using four imaging modalities: structural images (T1w and T2w), resting-state fMRI (rfMRI), task-fMRI (tfMRI), and high angular resolution diffusion imaging (dMRI). Behavioral and other individual subject measure data (both NIH Toolbox and non-Toolbox measures) is available on all subjects. MEG data and 7T MR data is available for a subset of subjects (twin pairs). The Open Access Dataset includes imaging data and most behavioral data. All details in the imaging protocols can be found at study webpage (https://humanconnectome.org/study/hcp-young-adult/document/1200-subjects-data-release/)

